# High frequency post-pause word choices and task-dependent speech behavior characterize connected speech in individuals with mild cognitive impairment

**DOI:** 10.1101/2024.02.25.24303329

**Authors:** Michael J. Kleiman, James E. Galvin

## Abstract

**Background:** Alzheimer’s disease (AD) is characterized by progressive cognitive decline, including impairments in speech production and fluency. Mild cognitive impairment (MCI), a prodrome of AD, has also been linked with changes in speech behavior but to a more subtle degree.

**Objective:** This study aimed to investigate whether speech behavior immediately following both filled and unfilled pauses (post-pause speech behavior) differs between individuals with MCI and healthy controls (HCs), and how these differences are influenced by the cognitive demands of various speech tasks.

**Methods:** Transcribed speech samples were analyzed from both groups across different tasks, including immediate and delayed narrative recall, picture descriptions, and free responses. Key metrics including lexical and syntactic complexity, lexical frequency and diversity, and part of speech usage, both overall and post-pause, were examined.

**Results:** Significant differences in pause usage were observed between groups, with a higher incidence and longer latencies following these pauses in the MCI group. Lexical frequency following filled pauses was higher among MCI participants in the free response task but not in other tasks, potentially due to the relative cognitive load of the tasks. The immediate recall task was most useful at differentiating between groups. Predictive analyses utilizing random forest classifiers demonstrated high specificity in using speech behavior metrics to differentiate between MCI and HCs.

**Conclusions:** Speech behavior following pauses differs between MCI participants and healthy controls, with these differences being influenced by the cognitive demands of the speech tasks. These post-pause speech metrics can be easily integrated into existing speech analysis paradigms.

## INTRODUCTION

Alzheimer’s disease (AD) is characterized by progressive degradation of cognitive abilities that interfere with everyday functioning. While episodic memory is often impacted early, other domains also may show early deficits. The regions associated with language and speech production are often among the earliest affected [1,2]. Recent studies have provided insight into how AD affects speech production, including the increased use of more common words [3,4], a potentially compensatory mechanism of declining executive functioning, and less content-filled language [5] which may reflect degradation of semantic memory. Individuals with AD have also been found to exhibit increased pauses during speech [5–7], potentially indicative of difficulties in planning and increased lexical search.

Mild cognitive impairment (MCI), a prodromal stage of AD [8], has been associated with the degradation of more subtle measures of speech behavior in its earliest stages. Measures of speech fluency, including rate of speech and syllable count, have been shown to significantly degrade in MCI [9,10]. These are generally measures of connected speech performance; a form of speech in which each individual word is “connected” lexically to the next, as in a passage or narrative. One of the most common methods of obtaining a sample of connected speech is directing the participant to describe a scene or image, such as the Cookie Theft image [11], which enables participants to produce a descriptive sample of speech that is also restricted to a particular topic to promote scoring ability. A more open-ended approach is to simply ask the participant to freely answer a given question, such as “what did you do yesterday?”, which prompts the production of more natural and conversational speech behavior. When scoring episodic memory is more important than capturing naturalistic speech, a paragraph recall task may also be used, as in the Wechler Logical Memory or Craft Story tests [12,13]. In these tasks, a story is read or shown to a participant who is then asked to recall as many details about the story as possible both immediately following the story, as well as after a delay.

In connected speech tasks, some studies find that MCI is associated with more common, less complex language [14–16], which may be related to a delay in accessing lexical information [17] resulting in a tendency to use high frequency (more common) words, however some other studies do not observe this increase in lexical frequency within MCI participants [18]. This trend towards common language is most visible in more severe forms of dementia, including primary progressive aphasia [19,20] and AD [3,4], suggesting that while the limited findings of increased lexical frequency due to MCI may be reflective of markers of future decline, the heterogeneity of the disorder may make it more difficult for these markers to be detected.

Pauses, or periods between clauses where a speaker hesitates or stops speaking, have also been shown to be affected in MCI. Pauses are characterized as either filled or unfilled. While unfilled pauses consist of extended pauses between words without any vocalization, often defined at greater than 250ms [21] but occasionally at a lower threshold or with no minimum threshold [22], filled pauses include utterances such as “uh”, “um”, and “er”, and are often used when a speaker is attempting to think of the next word or phrase, marking a region in which the speaker is engaging in word retrieval or language planning [23]. The usage of different “forms” of filled pauses have also been shown to vary in their usage; “uh” is typically used to indicate a short pause, while “um” is more often followed by longer pauses [24]. The frequency of these pause forms differ based on the task administered [25] which matches findings showing that pause usage in MCI changes based on the type of assessment; MCI is associated with increases in only filled pauses in free-response tasks [26,27] and either only unfilled pauses [27] or both filled and unfilled pauses [28] in narrative recall tasks.

Given that pauses are often produced during word searching behavior [23] and impaired word searching behavior has been found to lead to increased lexical frequency in MCI [14,16,17], we hypothesize that words immediately following pauses (“post-pause” behavior) are more likely to be higher frequency and syntactically simple in MCI participants compared to post-pause word choices in cognitively-intact healthy controls (HCs). Further, previous studies have shown that speech behavior is variably affected by task [16,27,29], as well as by the cognitive demands involved in performing that task [10]. As a result, we expect that differences in pauses and post-pause word choices will differ based on the degree of task demands for each of the three tasks that we will examine: narrative recall (immediate and delayed), picture descriptions, and free response. In particular, both the immediate and delayed narrative recall tasks which involve participants engaging in episodic memory recall are more difficult and require more attention and focus compared to tasks with lesser demands such as describing a picture or simply answering a question. We thus secondarily hypothesize that not all pauses are lexically-driven; more difficult tasks, especially the delayed narrative recall task, may increase pauses but without affecting post-pause lexical frequency, and vice versa.

## METHODS

### Participants

53 participants (39 HCs and 14 participants with MCI) were selected from the Healthy Brain Initiative, a longitudinal study of brain health and cognition at the University of Miami Comprehensive Center for Brain Health [30] based on their consensus clinical diagnosis (no cognitive impairment or mild cognitive impairment) and their age (greater than 60 years old). A full description of the study protocol is described elsewhere [30], but in-brief every participant undergoes identical annual comprehensive clinical, cognitive, functional, and behavioral assessments, including the Clinical Dementia Rating (CDR) [31], a complete physical and neurological examination, and neuropsychological test battery from the Uniform Data Set [32], supplemented with additional tests. Healthy controls are defined as individuals with a global CDR of 0, unimpaired activities of daily living, and normal age- and education-normed neuropsychological test performance. Individuals with MCI were defined as a global CDR 0.5, unimpaired activities of daily living, and neuropsychological test performance in at least 1 domain greater than 1.5 standard deviations below the norm. Imaging and plasma biomarker collection is ongoing so only clinical diagnoses were considered here. Exclusion criteria for this study included presence of clinically diagnosed aphasia, schizophrenia, generalized anxiety disorder or major depressive disorder, non-Alzheimer’s dementias, other neurological disorders including Parkinson’s disease or stroke, and cancer diagnosed within the past five years has not been determined to be in remission by a physician. All participants identified English as their primary language. All protocols were approved by the Institutional Review Board at the University of Miami Miller School of Medicine.

### Equipment and Materials

Participants were seated in a quiet area with minimal visual and auditory distractions in front of a 24” computer monitor attached to a Windows 10 machine. Participants interacted with the machine using custom-developed software written in Python and C, as well as a backlit keypad for indicating intention (finished with response, need help, etc.) and a volume knob. Participants wore Sony WH1000XM4 noise-cancelling headphones to further reduce auditory distractions. The software displayed visual and auditory stimuli for all tasks as well as written and audio instructions. Participants were given the opportunity to indicate whether they did not understand instructions and needed further explanation, after which an experimenter would assist them.

### Procedure

#### Speech Tasks

Three speech-focused tasks were administered to participants: narrative recall with delay, picture description, and a spontaneous free response. Participant audio was recorded using a hypercardioid microphone to minimize background noise.

The **narrative recall (NR)** task uses a custom-developed story (“Maddy the dog”) structured to be similar to the Craft Story 21 and Weschler Logical Memory narratives, with 25 content units of varying complexity [33] and developed to be used alongside the Craft Story 21 [13] which is administered to all participants in a separate visit as part of their longitudinal evaluations. This story differs from traditional narrative assessment in that it is both verbally read and visually presented to participants, promoting stronger multimodal encoding than simple auditory presentation. Additionally, the delayed portion was collected between fifteen and twenty minutes after initial presentation of the story, in contrast to the traditional 30 minutes, to both streamline data collection and explore differences between the already collected Craft Story delay. During this delay, additional speech tasks were administered. Participants were given two minutes for both the immediate and delayed responses.

**Picture descriptions (PD)** for a black and white version of the Modern Cookie Theft image [34] were collected from each participant, chosen as it is a highly validated and commonly-used stimulus for eliciting descriptive spontaneous speech. The updated image, as opposed to the original line-drawing version [11], is a more modern depiction of a kitchen scene which avoids stereotypical gender roles, adds additional content units, and incorporates more robust shading and coloring as opposed to the flat line drawings of the original which may aid participants in visually exploring the scene [35]. This updated version has been shown to elicit more detailed descriptions even under time constraints [36]. We further transformed this version by desaturating the image while still maintaining high contrast, as preliminary study revealed that many participants overly relied on color-based descriptions and less on describing action. Participants are given ninety seconds to describe each image, with a visual indicator at the 80-second mark. Additionally, if they choose to end their description before the time limit, they are asked “is there anything else?”.

**Spontaneous free responses (FR)** to the prompt “Describe your typical morning routine” were also collected. Participants were given no additional prompting, with their responses recorded until they indicated completion or until 90 seconds had passed.

#### Transcripts

Audio recordings from each task were processed using *OpenAI Whisper*’s Large-v2 model [37], a speech-to-text model that generates transcripts at higher accuracy than many other free and paid options and is capable of being run on in-house hardware. Each automatically generated transcript was then manually corrected as needed by trained research staff. The majority of corrections included the removal of experimenter instructions, hallucinations including “The end” and “Thanks for watching”, and obscured words due to high ambient noise, however the rate of corrections including the word error rate was not tracked. Generated transcripts included start and stop times in seconds for each word as well as confidence for each predicted word which helped highlight areas of potentially necessary correction.

#### Audio Preprocessing

All audio files were preprocessed prior to speech-to-text transcription. Leading and trailing silence was removed using *PyDub* [38], and the end of a file was marked using a chime to encourage *OpenAI Whisper* to end sentences appropriately when subjects ran out of time and the audio file ended abruptly. If a chime is not used, hallucinations where *Whisper* finished sentences were common, including erroneously completed sentences with predicted text or an ending message (e.g., “The end.”) Background noise, including room tone and non-target extraneous speech (e.g., voices from hallway) were removed using *PySoX* [39] to further improve speech-to-text accuracy as well as to facilitate audiometric analysis.

### Analysis

#### Analysis of Speech Transcripts

After speech responses were transcribed and validated, the transcripts were parsed using a Python script incorporating multiple NLP packages, including *SpaCy* for tokenization, lemmatization, part of speech analysis, and general NLP analyses [40], *wordfreq* for examining lexical frequency using its own proprietary formula [41], and *textdescriptives* for generating a wide range of statistics including entropy, perplexity, sentence count and length, syllable count, and reading ease statistics such as Flesch Kincaid grade level, Gunning-Fog, and the Coleman Liau Index [42,43]. Custom scripts were also written that generate SUBTLEX rankings [44] for determining lexical frequency, mean Yngve depth for examining passage complexity [45], and type token ratios and derivatives (MATTR or “moving average type token ratio” [46] and MTLD or “measure of textual diversity” [47]) for lexical diversity. Distinctions are made for which corpus (wordfreq or SUBTLEX) was utilized when examining lexical frequency, as each produces slightly different results due to using different word lists.

Pauses were defined as either filled or unfilled, with unfilled pauses defined as pauses lasting greater than 250 milliseconds between words, based on literature examining pauses in normal conversational speech [22] as well as patients with aphasia [21]. Filled pauses were defined as the presence of “um”, “uh”, and “er”, chosen based on their frequency in English speech [48]. Both filled and unfilled pauses were calculated automatically using Python scripts; unfilled pauses were marked when durations greater than 250ms were detected between the end of the previous word and the start of the current word, as determined by timestamps generated by OpenAI Whisper, and filled pauses were determined by tokens containing a filler word. The four words immediately following the pause were marked as “post-pause” words, and statistics were generated for both the average of all four words and only the first word following the pause. These pause and post-pause statistics included total counts and durations of pauses, latencies, syllable counts, SUBTLEX rankings, *wordfreq* frequencies, and part of speech frequencies. Pause duration was defined as the total time in milliseconds between the start of the pause (end of the previous word) and the beginning of the following word. For filled pauses, latency was defined as the time from the end of the utterance of the filler word to the beginning of the following word, and for unfilled pauses latency was only used in the context of calculating the latencies of all pauses in which case pause duration was used.

#### Statistical and Predictive Analyses

Transcripts were compared between groups for both individual and combined tasks using the *pingouin v0.5.3* [49] Python package. Analyses of covariance (ANCOVA) with age as a covariate were performed for all measures where assumptions of normality and homoscedasticity were met, and Kruskal-Wallis Rank Sum test when assumptions were not met and age was not significantly different between groups as determined by linear regression. In cases where assumptions were not met and age was a significant factor, comparisons were not considered. Statistical logistic regressions were also used to determine the combined effects of multiple variables, especially when isolating the effects of post-pause metrics alone. Pearson’s correlations were used to compare speech metrics identified as significantly different by ANOVAs with cognitive and neuropsychological assessments including the Montreal Cognitive Assessment (MoCA) [50], the Cognivue total [51], number span backwards, the Trail Making Test B, Hopkins Verbal Learning Task immediate and delayed [52], semantic verbal fluency (animals), and the Number Symbol Coding Task [53]. Verbal IQ assessed by the Test of Premorbid Functioning as well as the Vulnerability Index [54] and the Resilience Index [55] were also included in correlational analyses with the identified speech metrics. See Besser et al. [30] for details on the administration and examination of these assessments.

Predictive analyses, including random forest classifiers, gradient boosted machines, logistic regressions, and support vector classifiers, were performed to identify the predictive power of post-pause and general speech metrics to determine binary impairment status (impaired vs. not impaired). All analyses were cross-validated using a repeated stratified K-folds procedure (3-fold, 3-repeat), which resulted in nine combinations of train/test sets for better generalizability of model results. Outputs were evaluated using sensitivity, specificity, F1 score, and area under the ROC curve (AUC). The *LightGBM v4.2.0* [56] Python package’s implementation was used for gradient boosted machines, and all other predictive analyses and methods were performed using *scikit-learn v1.3.2* [57].

Two sets of features were used in our models; one including all statistically significant features, and another including only statistically significant post-pause metrics (**Table 3**). Only individual task metrics were used; when significant results for were found in the mixed ANOVA containing all tasks, only the task with the highest Bayes index in the post-hoc was used in an effort to minimize the negative effect of multicollinearity on model results. Additionally, feature selection was performed using the *BorutaSHAP v1.1* [58] selection algorithm using a random forest classifier (*scikit-learn v1.3.0* [57]) as its base model. One-fourth (25%) of the stratified training data was held for use in the feature selection process, to avoid leakage and subsequent overfitting in the model training phase. *Optuna v3.5.0* [59] selected optimal hyperparameters for each model, leveraging its implementation of define-by-run dynamic parameter search spaces and efficient strategies for pruning, using the same 25% of stratified training data in the feature selection step.

## RESULTS

### Sample Characteristics

There was a significant difference in age between the two groups (*p* = .003), resulting in age being included as a covariate in all following comparisons (**Table 1**). No differences between groups were found for gender, years of education, race or ethnicity, vulnerability [54], or resilience [55]. Significant differences in cognitive tasks were observed between groups, consistent with a classification of MCI.

**Table 1.** Subject characteristics.

|  | Healthy Controls | Mild Cognitive Impairment | p-val |
| --- | --- | --- | --- |
| Age | 68.28 (7.21) | 76.64 (8.90) | **0.000** |
| % Female | 0.72 (0.45) | 0.64 (0.50) | 0.478 |
| Years of Education | 16.43 (2.65) | 17.00 (3.22) | 0.920 |
| % Non-White | 0.11 (0.31) | 0.21 (0.43) | 0.159 |
| % Hispanic | 0.12 (0.33) | 0.21 (0.43) | 0.130 |
| MoCA Total | 27.05 (2.26) | 23.29 (2.73) | **0.000** |
| FAQ Total | 0.14 (0.45) | 0.18 (0.60) | 0.750 |
| Numbers Backwards | 5.37 (1.33) | 3.86 (0.86) | **0.000** |
| Trailmaking B | 68.11 (24.34) | 188.08 (241.85) | **0.000** |
| Categorical Fluency | 21.21 (5.19) | 16.50 (4.03) | **0.027** |
| HVLT Immediate | 25.32 (3.80) | 20.07 (5.05) | **0.004** |
| HVLT Delayed | 9.52 (1.84) | 6.21 (3.89) | **0.001** |
| Craft Story Immediate | 21.20 (5.68) | 18.93 (6.20) | 0.198 |
| Craft Story Delayed | 19.17 (5.56) | 14.50 (8.30) | **0.020** |
| Verbal IQ | 116.33 (9.44) | 111.50 (12.06) | **0.008** |
| Vulnerability Index | 6.55 (2.02) | 8.64 (2.73) | 0.078 |
| Resilience Index | 183.33 (33.06) | 172.54 (32.30) | 0.326 |
| Number Symbol Coding Test | 45.89 (9.16) | 31.36 (6.49) | **0.000** |

All comparisons performed using ANCOVA with age as a covariate, except age which used ANOVA.
MoCA = Montreal Cognitive Assessment; FAQ = Functional Activities Questionnaire; HVLT = Hopkins Verbal Learning Test
Bold = significant

### Common Findings in All Tasks

When examining all tasks using a Mixed ANOVA, with task as the within-subjects variable and impairment status as the between-subjects variable, differences were found between the HC and MCI groups. There were significantly more filled pauses (“um”, “uh”, and “er”) in the MCI group (4.21 ± 4.42) compared to the HC group (2.47 ± 2.71), *p*=.006, especially containing “uh” (2.75±3.53 MCI vs 1.11±1.78 HC; χ^2^(1)=13.31, p<.001), with latencies between all pauses and the following word increasing in the MCI group (1.22±0.35 MCI vs 1.11±0.33 HC; χ^2^(1)=9.31, *p*=.002), **Figure 1**.

**Figure 1.**
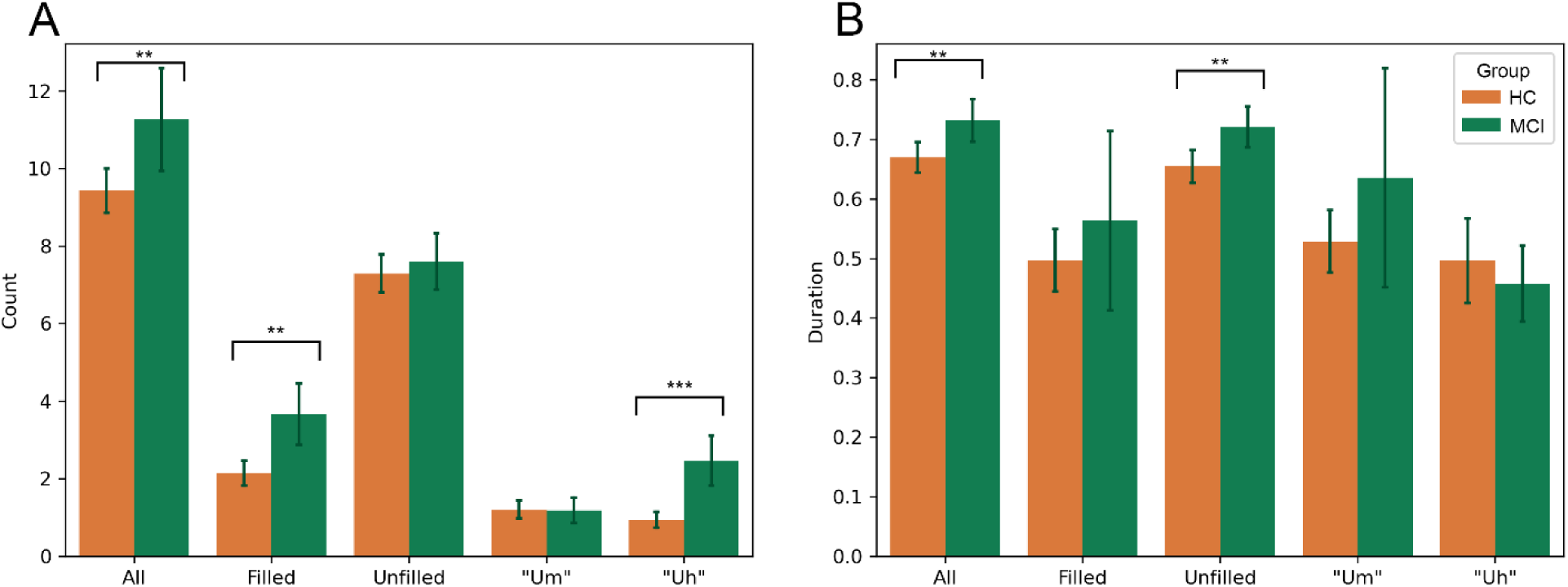
Count and duration of pause types separated by impairment status. (A) Total number of pauses separated by type of pause (all pauses, filled, unfilled, filled with “um”, filled with “uh”) and by impairment status (HC in orange vs MCI in green). All tasks were included in these comparisons. Significant differences between impairment status can be observed when examining All pauses, Filled pauses, and “Uh” pauses, all of which increased in the MCI group, with no differences found for unfilled and “um” pauses when all tasks were included. (B) Total duration in seconds separated by type of pause and impairment status. Unfilled pauses and All pauses were found to significantly increase in the MCI group. Significance is indicated using asterisks, with *** = *p* < .001, ** = *p* < .01, * = *p* < .05.

Correlational analyses with cognitive and neuropsychological assessments revealed that total word count was often mildly or moderately negatively correlated with decreased cognitive functioning, including the MoCA (r=.190, *p*=.002), Cognivue (r=.168, *p*=.009), trailmaking test B (r=-.214, *p*<.001), semantic verbal fluency (r=.198, *p*=.002), and the Number Symbol Coding Task (r=.194, *p*=.002), as well as verbal IQ as assessed by the Test of Premorbid Functioning (r=.269, *p*<.001) and the Resilience Index (r=.250, *p*<.001). Despite “um” pause counts alone not being significantly different between groups, moderate negative correlations were found with the MoCA (r=-.261, *p*<.001), Cognivue (r=-.308, *p*<.001), Number Symbol Coding (r=-.232, *p*<.001), and semantic verbal fluency (r=-.196, *p*=.003) tests, in addition to the Resilience Index (r=-.251, *p*<.001); a mild positive correlation was also found with the Vulnerability Index (r=.179, *p*=.005).

For post-pause metrics, the Kruskal-Wallis non-parametric ANOVA test was required due to many participants not producing pauses at all in some tasks, resulting in unequal variances between groups. Lexical frequency was significantly higher following a filled pause in MCI participants, supporting our hypotheses; *wordfreq* rankings following “uh” fillers (.0087±.013 MCI vs .0058±.012 HC; χ^2^(1)=7.31, *p*=.007) were significantly different. Latencies following “um” fillers were also significantly higher in MCI participants (1.83 sec ± 0.73 MCI vs 1.33 sec ± 0.56 HC; χ^2^(1)=17.36, *p*<.001). Mean latency following pause filled with “um” was negatively correlated with the Cognivue (r=-.335, *p*<.001), Hopkins Verbal Learning immediate (r=-.244, *p*=.005) and delayed (r=-.259, *p*=.003), semantic verbal fluency (r=-.285, *p*=.002), and the Number Symbol Coding task (r=-.323, *p*<.001), in addition to a positive correlation with Trailmaking B (r=.312, *p*<.001).

### Task Comparisons

All differences between tasks described below exhibited a significant ANOVA or pairwise t-test within-subjects score, with an alpha set at .015.

#### Picture description

In the PD task, the mean Yngve depth was shallower in MCI participants (3.36±0.67 levels) than for HC participants (3.98±0.86 levels), *p*=.012, however no significant correlations with cognitive assessments were identified. The Coleman-Liau Index, a measure of readability, also decreased in MCI participants (3.65±1.45 MCI vs 4.74±0.88 HC; *p*=.008) (**Table 2.c**), findings supported by a moderate correlation with the MoCA total (r=.395, *p*=.001). Total pause count was moderately negatively correlated with the Cognivue (r=-.405, *p*=.001) in this task, and the mean *wordfreq* lexical frequency was also negatively correlated with the MoCA total (-.338, *p*=.007). Other significant correlations are found in **Figure 2**.

**Figure 2.**
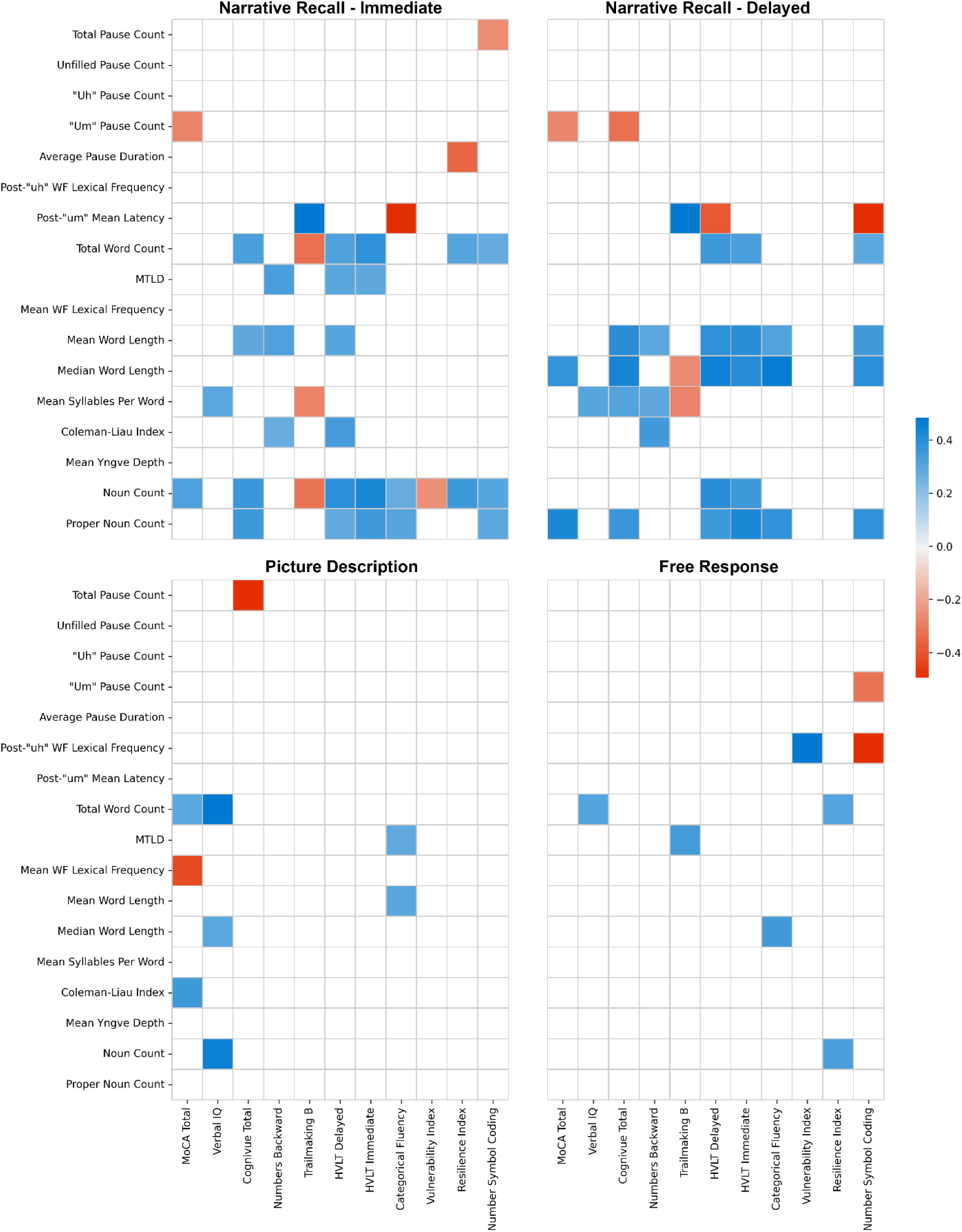
Correlation matrix using Pearson’s correlations to compare significant speech metrics with cognitive and neuropsychological assessments for each of the four tasks. Spaces without color are non-significant determined by a *p-*value above 0.015.

#### Narrative recall

In both narrative recall tasks, filled pauses significantly increased in MCI participants (3.23±3.97 pauses MCI vs 1.58±2.28 HC; χ^2^(1)=7.37, *p*=.007), however this significance did not appear in pairwise comparisons (**Table 2.a, 2.b**). Latencies following “um” fillers increased in MCI participants in both NR tasks (1.82±0.72 sec MCI vs 1.25±0.49 sec HC; χ^2^(1)=8.43, *p*=.004) and were significantly correlated with measures of cognition (**Figure 2**), however post-hoc analyses revealed that only the delayed NR task showed significance between groups (**Table 2.b**).

**Table 2.a.** Speech measures for the Immediate Narrative Recall task.

| Speech measure | HC | MCI | Stat (df = 60) | p-val | Test |
| --- | --- | --- | --- | --- | --- |
| Total Pause Count | 7.325 (3.422) | 9.133 (4.809) | 2.138 | 0.144 | KW |
| Unfilled Pause Count | 5.975 (2.824) | 6.533 (2.588) | 1.179 | 0.278 | KW |
| “Uh” Pause Count | 0.707 (1.470) | 1.333 (1.633) | 3.167 | 0.075 | KW |
| “Um” Pause Count | 0.610 (1.321) | 2.200 (3.570) | 2.509 | 0.113 | KW |
| Post-“uh” WF Lexical frequency | 0.008 (0.016) | 0.004 (0.008) | 0.029 | 0.866 | KW |
| Post-“um” Mean Latency | 1.395 (0.471) | 1.762 (0.721) | 1.970 | 0.160 | KW |
| Total Word Count | 84.378 (26.173) | 70.059 (24.517) | 3.818 | 0.055 | ANOVA |
| MTLD | 50.422 (15.642) | 38.791 (16.528) | 6.616 | **0.013** | ANOVA |
| Mean WF Lexical frequency | 0.006 (0.001) | 0.007 (0.002) | 0.557 | 0.458 | ANOVA |
| Mean Word Length | 4.158 (0.239) | 3.944 (0.258) | 9.449 | **0.003** | ANOVA |
| Median Word Length | 3.967 (0.165) | 3.735 (0.437) | 7.772 | 0.005<sup>x</sup> | KW |
| Mean Syllables per Word | 1.156 (0.046) | 1.117 (0.055) | 7.980 | **0.006** | ANOVA |
| Coleman-Liau Index | 6.737 (1.670) | 5.271 (2.306) | 7.660 | **0.007** | ANOVA |
| Mean Yngve Depth | 4.785 (1.450) | 4.739 (1.733) | 0.011 | 0.917 | ANOVA |
| Noun Count | 12.733 (3.774) | 9.824 (3.226) | 7.902 | **0.007** | ANOVA |
| Proper Noun Count | 5.244 (2.542) | 2.706 (2.392) | 12.694 | **0.001** | ANOVA |

KW = Kruskal-Wallis; ANOVA = Analysis of Variance; HC = Healthy Control; MCI = Mild cognitive impairment; MTLD = measure of textual lexical diversity; X = Significant age effects; WF = *wordfreq* lexical frequency
Bold = significant

**Table 2.b.** Speech measures for the Delayed Narrative Recall task.

| Speech measure | HC | MCI | F (df = 60) | p-val | Test |
| --- | --- | --- | --- | --- | --- |
| Total Pause Count | 9.375 (5.138) | 9.000 (7.774) | 2.168 | 0.141 | KW |
| Unfilled Pause Count | 7.500 (5.208) | 6.333 (5.080) | 2.34 | 0.126 | KW |
| “Uh” Pause Count | 1.140 (1.656) | 1.062 (1.611) | 0.123 | 0.726 | KW |
| “Um” Pause Count | 0.698 (1.264) | 2.062 (2.720) | 4.682 | 0.030 | KW |
| Post-“uh” WF Lexical frequency | 0.003 (0.003) | 0.005 (0.004) | 1.231 | 0.267 | KW |
| Post-“um” Mean Latency | 1.174 (0.500) | 1.886 (0.766) | 5.740 | **0.015** | KW |
| Total Word Count | 96.065 (35.668) | 79.750 (35.422) | 2.492 | 0.120 | ANOVA |
| MTLD | 50.375 (23.418) | 45.202 (14.668) | 0.683 | 0.412 | ANOVA |
| Mean WF Lexical frequency | 0.007 (0.002) | 0.007 (0.001) | 0.146 | 0.704 | ANOVA |
| Mean Word Length | 4.109 (0.334) | 3.889 (0.226) | 5.954 | 0.018 | ANOVA |
| Median Word Length | 3.902 (0.467) | 3.500 (0.483) | 9.827 | **0.002** | KW |
| Mean Syllables per Word | 1.168 (0.054) | 1.137 (0.060) | 3.747 | 0.058 | ANOVA |
| Coleman-Liau Index | 5.905 (2.242) | 5.128 (2.519) | 1.339 | 0.252 | ANOVA |
| Mean Yngve Depth | 4.639 (1.404) | 5.156 (1.741) | 1.420 | 0.238 | ANOVA |
| Noun Count | 15.848 (5.692) | 12.688 (6.488) | 3.405 | 0.070 | ANOVA |
| Proper Noun Count | 4.891 (2.915) | 1.938 (1.692) | 13.026 | **0.000** | ANOVA |

KW = Kruskal-Wallis; ANOVA = Analysis of Variance; HC = Healthy Control; MCI = Mild cognitive impairment; MTLD = measure of textual lexical diversity; X = Significant age effects; WF = *wordfreq* lexical frequency
Bold = significant

Mean total word counts decreased in both immediate and delayed NR tasks (74.75±30.21 words in MCI vs 92.88±31.95 words in HC; *p*=.002), with lexical diversity measured by MTLD found to decrease in only the immediate NR task (38.79±16.53 MCI vs 50.42±15.64 HC; *p*=.013). Also in the immediate NR task, the Coleman Liau Index decreased from 6.74±1.67 in HC to 5.27±2.31 in MCI, *p*=.007. Mean word lengths (3.94±0.26 characters MCI vs 4.16±0.24 characters HC; *p*=.003) in the immediate NR task also decreased in MCI participants, while median word length decreased in the delayed NR task (3.50±0.48 MCI vs 3.90±0.47 HC; p=.002). The number of syllables per word in the immediate NR task was also found to decrease for MCI participants (1.12±0.06 syllables MCI vs 1.16±0.05 syllables HC; *p*=.006). Fewer nouns were used by MCI participants in the immediate NR task (9.82±3.23 nouns MCI vs 12.73±3.77 nouns HC; *p*=.007). Further, proper nouns such as names were significantly less common for MCI participants in both the immediate NR (2.71±2.39 words MCI vs 5.24±2.54 words HC; *p*=.001) and delayed NR tasks (1.94±1.69 words MCI vs 4.89±2.92 words HC; *p*<.001), findings supported by significant correlations for both nouns and proper nouns with most cognitive and neuropsychological assessments in these tasks (**Figure 2**).

#### Free response

Word frequencies as calculated by *wordfreq* were significantly more common in MCI participants after “uh” fillers in this task (.0086±.0078 MCI vs .0025±.0027 HC; *p*=.005) (**Table 2.d**), but not in other tasks (**Figure 3**). Supporting this, strong correlations were found between the Number Symbol Coding Task (r=-.496, *p*=.002) and the Vulnerability Index (r=.483, *p*=.003). Additionally, adverb usage significantly decreased following unfilled pauses in MCI participants (1.8% ± 3.9% adverbs post-pause) compared to HCs (16.4% ± 20.8% adverbs post-pause), χ^2^(1)=8.89, *p*=.003.

**Figure 3.**
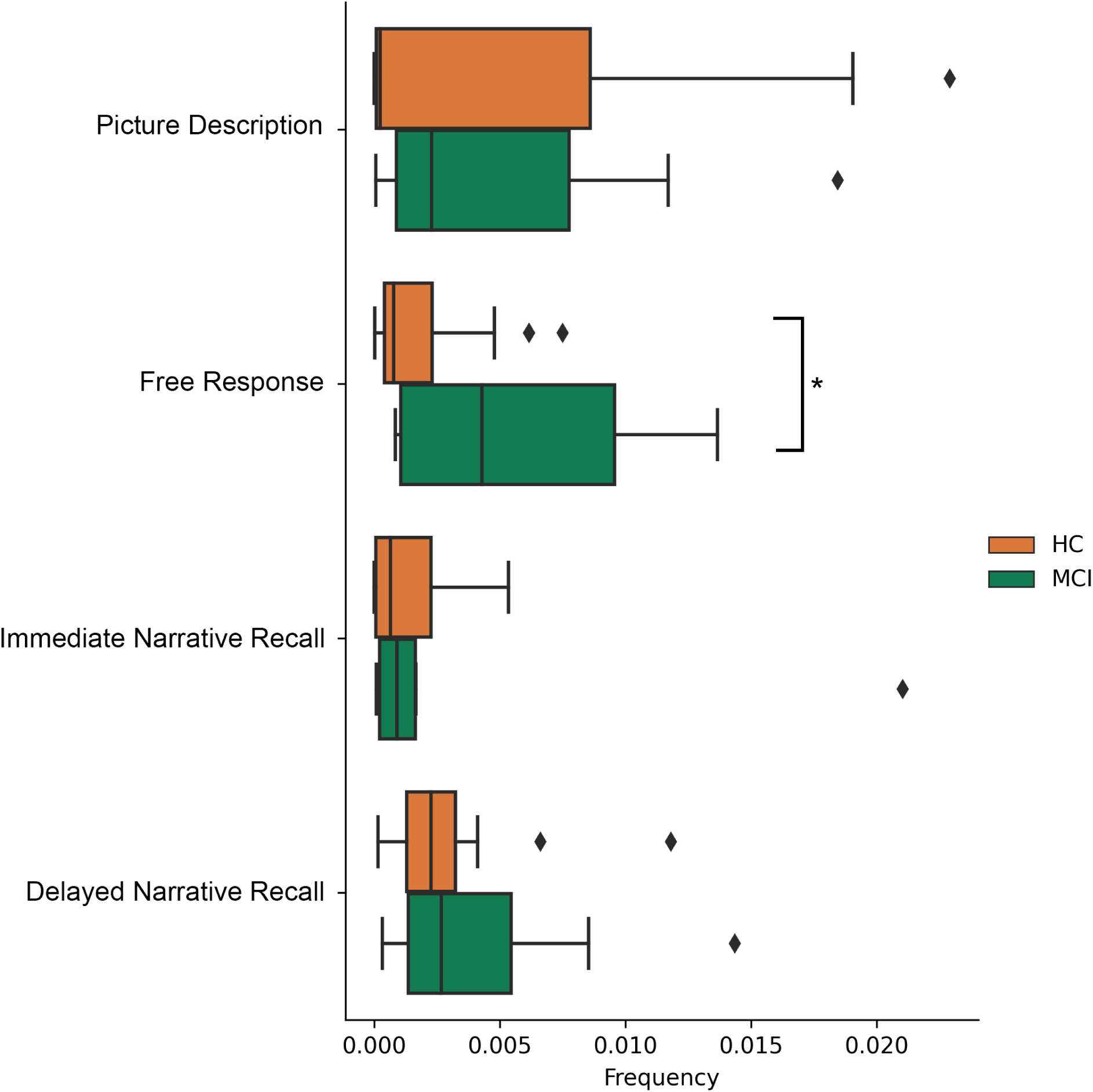
Task comparisons of post-“uh” pause lexical frequency rankings. This measure was used for comparison to highlight differences in pause and post-pause behavior between tasks. In both narrative recall tasks, all groups tended to be searching for comparatively uncommon words following “uh” pauses, while in the picture description task all groups searched for more common words. Only in the free response task were there differences between impairment status, with healthy controls (HC) searching only for more uncommon words and participants with mild cognitive impairment (MCI) producing words of more variable frequency following “uh” pauses.

**Table 2.c.** Speech measures for the Picture Description task.

| Speech measure | HC | MCI | F (df = 60) | p-val | Test |
| --- | --- | --- | --- | --- | --- |
| Total Pause Count | 11.561 (5.473) | 14.125 (6.195) | 0.397 | 0.531 | ANCOVA |
| Unfilled Pause Count | 8.854 (4.709) | 10.500 (5.304) | 0.018 | 0.893 | ANCOVA |
| “Uh” Pause Count | 1.178 (1.482) | 1.471 (1.068) | 0.553 | 0.460 | ANOVA |
| “Um” Pause Count | 1.400 (1.959) | 2.588 (2.476) | 3.916 | 0.052 | ANOVA |
| Post-“uh” WF Lexical frequency | 0.010 (0.016) | 0.014 (0.019) | 1.720 | 0.190 | KW |
| Post-“um” Mean Latency | 1.498 (0.720) | 1.912 (0.752) | 2.658 | 0.103 | KW |
| Total Word Count | 134.356 (55.009) | 114.176 (43.805) | 1.840 | 0.180 | ANOVA |
| MTLD | 38.641 (11.760) | 33.141 (12.431) | 2.617 | 0.111 | ANOVA |
| Mean WF Lexical frequency | 0.010 (0.002) | 0.010 (0.002) | 0.934 | 0.338 | ANOVA |
| Mean Word Length | 3.981 (0.143) | 3.843 (0.179) | 4.638 | 0.035<sup>x</sup> | ANCOVA |
| Median Word Length | 3.356 (0.472) | 3.000 (0.000) | 8.613 | **0.003** | KW |
| Mean Syllables per Word | 1.217 (0.040) | 1.191 (0.053) | 0.337 | 0.564<sup>x</sup> | ANCOVA |
| Coleman-Liau Index | 4.740 (0.884) | 3.651 (1.448) | 7.069 | **0.008** | KW |
| Mean Yngve Depth | 3.983 (0.863) | 3.356 (0.668) | 4.913 | **0.012** | ANOVA |
| Noun Count | 32.933 (13.048) | 25.941 (9.959) | 3.987 | 0.05 | ANOVA |
| Proper Noun Count | 0.002 (0.006) | 0.001 (0.003) | 0.268 | 0.606 | ANOVA |

KW = Kruskal-Wallis; ANOVA = Analysis of Variance; HC = Healthy Control; MCI = Mild cognitive impairment; MTLD = measure of textual lexical diversity; X = Significant age effects; WF = *wordfreq* lexical frequency
Bold = significant

**Table 2.d.** Speech measures for the Free Response task.

| Speech measure | HC | MCI | F (df = 60) | p-val | Test |
| --- | --- | --- | --- | --- | --- |
| Total Pause Count | 12.049 (7.218) | 11.867 (7.170) | 0.007 | 0.934 | ANOVA |
| Unfilled Pause Count | 8.488 (5.437) | 8.000 (4.342) | 0.098 | 0.756 | ANOVA |
| “Uh” Pause Count | 1.932 (2.366) | 1.933 (1.981) | 0.105 | 0.746 | KW |
| “Um” Pause Count | 1.682 (2.197) | 4.200 (4.945) | 3.281 | 0.070<sup>x</sup> | KW |
| Post-“uh” WF Lexical frequency | 0.003 (0.003) | 0.009 (0.008) | 8.027 | **0.005** | KW |
| Post-“um” Mean Latency | 1.252 (0.391) | 1.731 (0.716) | 5.250 | 0.022 | KW |
| Total Word Count | 126.000 (71.155) | 119.938 (84.409) | 0.078 | 0.781 | ANOVA |
| MTLD | 42.294 (24.467) | 41.056 (21.867) | 0.032 | 0.859 | ANOVA |
| Mean WF Lexical frequency | 0.007 (0.001) | 0.007 (0.002) | 1.177 | 0.282 | ANOVA |
| Mean Word Length | 3.597 (0.260) | 3.572 (0.212) | 0.121 | 0.729 | ANOVA |
| Median Word Length | 3.130 (0.341) | 3.031 (0.125) | 0.000 | 0.992<sup>x</sup> | ANCOVA |
| Mean Syllables per Word | 1.152 (0.057) | 1.145 (0.043) | 0.207 | 0.650 | ANOVA |
| Coleman-Liau Index | 3.446 (1.797) | 3.530 (1.239) | 0.030 | 0.864 | ANOVA |
| Mean Yngve Depth | 4.867 (1.909) | 4.731 (1.369) | 0.068 | 0.795 | ANOVA |
| Noun Count | 24.130 (13.016) | 19.375 (11.949) | 1.649 | 0.204 | ANOVA |
| Proper Noun Count | 0.009 (0.016) | 0.012 (0.032) | 0.122 | 0.728 | ANOVA |

KW = Kruskal-Wallis; ANOVA = Analysis of Variance; HC = Healthy Control; MCI = Mild cognitive impairment; MTLD = measure of textual lexical diversity; X = Significant age effects; WF = *wordfreq* lexical frequency
Bold = significant

#### Controlling for individual cognitive resources

To rule out the effects of varying cognitive resources within groups, the Trailmaking B time (measure of attention), Number-Symbol Coding Task (measure of executive function), Hopkins verbal learning task delayed (measure of episodic memory), Animal Naming (measure of categorical verbal fluency), and the Test of Premorbid Functioning (measure of verbal IQ) were used as covariates for all significant findings above. No results were rendered non-significant after controlling for these measures.

### Predictive Analysis

When starting with all significant features, feature selection narrowed the field to 16 of the best-performing features: total pause count (immediate NR), total filled pause count (immediate NR), total “uh” count (immediate and delayed NR), mean *wordfreq* frequency post-“uh” (FR), mean latency post-“um” (delayed NR), word count (immediate NR), MTLD (immediate NR), mean *wordfreq* frequency (immediate NR), median word length (delayed NR) Coleman-Liau Index (PD & immediate NR), mean Yngve depth (PD), proper noun ratio (immediate NR), and noun count (delayed NR). Using this set of features and after performing hyperparameter optimization, the best performing model was the random forest classifier with a diagnostics odds ratio (DOR) of 13.52 (**Table 3**) and an AUC of 0.828 (**Figure 4)**.

**Figure 4.**
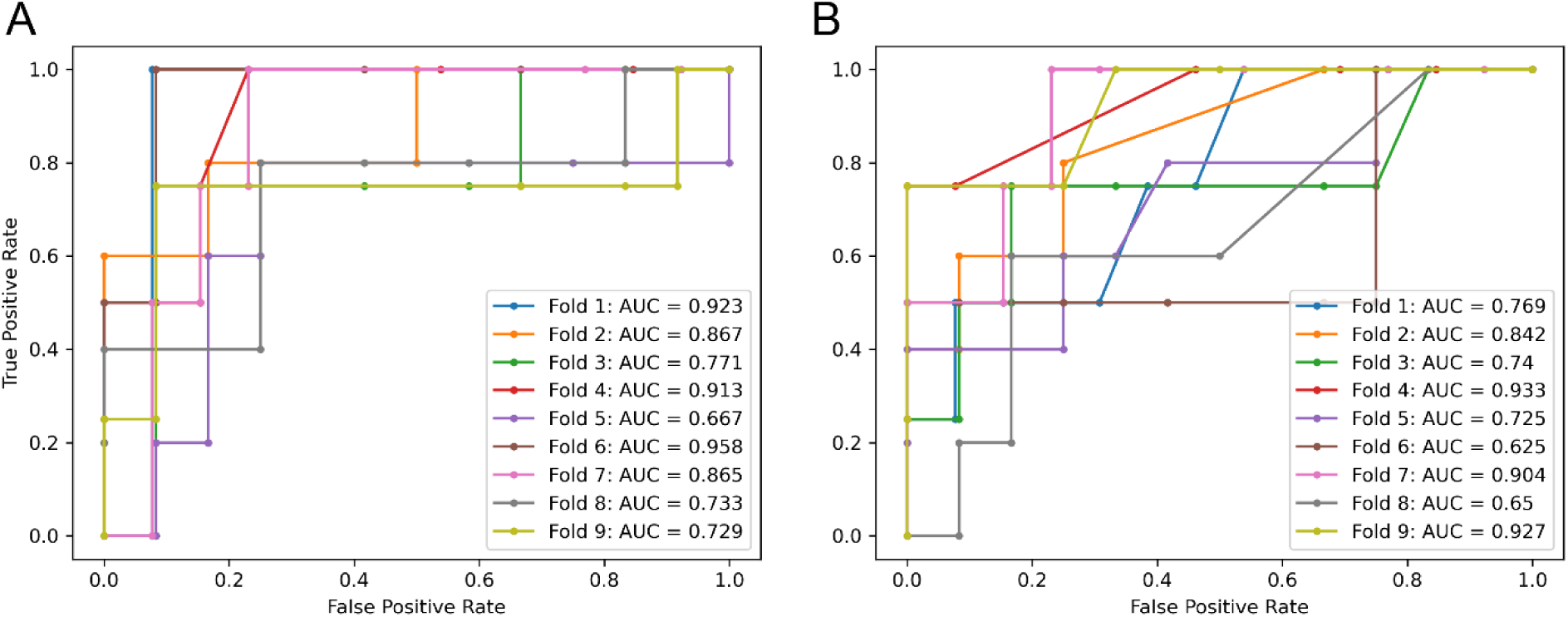
Areas under ROC curves for predictive models. (A) After performing feature selection on all available lexical features, 16 features were identified to best identify impairment status (HC vs MCI). Incorporating these into a repeated stratified 3-fold 3-repeat cross-validation procedure using *LightGBM* gradient boosted machines as the models generated a mean AUC of 0.828. AUCs for individual folds are depicted in multiple colors. (B) The best identified post-pause features (post-“uh” lexical frequency in free response and post-“um” latency in delayed narrative recall) were selected and used as sole features in a *LightGBM* gradient boosted machine, examined using repeated stratified 3-fold 3-repeat cross-validation. This parsimonious model performed similarly to the larger model with an AUC of 0.791. AUCs for individual folds are depicted in multiple colors.

**Table 3.** Measures of predictive ability for models utilizing speech behavior.

|  | **Sensitivity** | **Specificity** | **PPV** | **NPV** | **DOR** | **AUC** |
| --- | --- | --- | --- | --- | --- | --- |
| All Significant Features | 38.46% | 94.59% | 71.43% | 81.40% | 10.94 | 0.825 |
| Post-Pause Features | 41.03% | 93.70% | 69.57% | 81.89% | 10.33 | 0.791 |

**All Significant Features:** Total filled pauses (INR), Total “uh” pauses (INR & DNR), Mean *wordfreq* frequency post- “uh” (FR), mean latency post-“um” (DNR), word count (INR), MTLN (INR), mean *wordfreq* frequency (INR), median word length (DNR) Coleman-Liau Index (PD & INR), mean Yngve depth (PD), proper noun ratio (INR), noun count (DNR)
**Post-pause Features:** Mean *wordfreq* frequency post- “uh” (FR), mean latency post-“um” (DNR)
FR=“Free-response task”; INR=“Immediate Narrative Recall task”; DNR=“Delayed Narrative Recall task”; PD=“Picture Description task”; PPV=“Positive Predictive Value”; NPV=“Negative Predictive Value”; DOR=“Diagnostic Odds Ratio”; AUC=“Area under the ROC curve”

When using only the two most significant post-pause metrics (*wordfreq* lexical frequency following “uh” in the FR task, and latencies following “um” in the delayed NR task), the best performing model was again the random forest classifier with a DOR of 10.3 (**Table 3**) and an AUC of 0.791 (**Figure 4)**.

## DISCUSSION

In this study, we examined whether post-pause speech is affected due to cognitive impairment, and if task demands elicit differences in speech behavior between HC and MCI participants. We found significant differences between groups in *wordfreq* lexical frequency following filled pauses in the FR task, as well as increased latencies following “um” pauses in the NR tasks.

### Task-dependent pause production

MCI participants tended to use more common (high frequency) language following filled “uh” pauses, but only in the FR task. Our initial hypothesis that lexical frequency would decrease in MCI individuals due to increased word searching behavior may have underestimated the intervening effect of task demands. Just as different forms of filled pauses differ in their usage [24] it is possible that each form may be produced for different reasons as task conditions vary. Indeed, previous studies show that task difficulty and the associated increased cognitive load have been shown to affect speech production, including altering lexical diversity and syntactic complexity [60] as well as disfluency and pause usage [61]. The question in the FR task asked about the participant’s typical routine, which is a topic that is typically static, repetitive, and familiar, requiring few cognitive resources to recall effectively. Pauses produced in this context thus likely resemble typical conversational speech, and would match the documented usage of “uh” fillers as indicators of word-searching behavior [24,62,63]. As a result, cognitive impairment may play a larger role in disrupting this search, leading to greater differences between the HC and MCI groups. This is contrasted with the three other tasks, where required cognitive demands are greater to varying degrees. In these tasks, filled “uh” pauses may still be produced as indicators of searching behavior, but instead of lexical search being the driving force behind pause production as in the spontaneous speech task this searching behavior may be instead more directed towards searching for the correct answer (in NR tasks) or searching for a new object to describe (in the PD task). Thus, while post-filled pause lexical frequency may still be affected by cognitive impairment, the HC group may also use more common words as a result of the increased cognitive load [60], resulting in a less noticeable and thus non-significant difference between groups.

In a similar vein, the delayed NR task was arguably the most difficult task administered in this study, requiring participants to recall information presented after being distracted in the interim with other speech tasks while also being under a time limit [64]. Delayed recall activates broader neurological pathways, including activation of the left parahippocampal gyrus, the entorhinal and perirhinal cortices, and both the anterior and posterior hippocampal regions, contrasting with immediate recall which is handled primarily with short-term memory processes in the posterior hippocampus and dorsolateral prefrontal cortex [65,66]. This may explain why post-“um” latencies were significantly different between groups only in this task. “Um” fillers have been shown to signal longer delays than other pauses [24,63], and the delays decrease with skill [23] and increase with cognitive load [67]. Delayed narrative recall performance is known to be significantly affected due to cognitive impairment [68], and so MCI participants likely needed to utilize more cognitive resources to perform the task, leading to increased delays compared to the HC group.

While previous studies have shown significant differences between MCI and HC in numbers of pauses, both filled and unfilled [27,28] and especially using the “uh” utterance [69], our results did not show significant differences between the two groups in total count of pauses when examining the post-hoc pairwise tests for task comparison that were not also significantly different due to age effects. While our sample size is relatively limited, the observation of significant age effects may suggest that previous identification of increased numbers of pauses in these studies may be due to the MCI group being older than the HC group. Future examination into total pause counts should take age into account as a possible covariate.

### Other lexical differences between tasks

More significant differences between groups were found in the immediate NR task than any others, and when multiple tasks exhibited the same differences in a single measure the *Boruta* feature selection algorithm identified the immediate NR task as the most predictive. The MTLD measure of lexical diversity as well as the Coleman-Liau index both identify a tendency of MCI participants to use simpler language in this task, as does the finding of reduced mean word length and syllables per word. The high usefulness of the immediate NR task has been previously shown in prior work developing parsimonious assessment composites for detecting MCI [68], where it was determined to be one of the most useful metrics for identifying MCI among the neuropsychological tasks administered by ADNI. This may be due to this task requiring coordination of working memory and attention, two areas known to be degraded in MCI but not entirely disrupted.

Proper noun usage decreased in both the immediate and delayed NR tasks, indicating that while healthy participants are able to recall names and places, those with MCI are less likely to recall this information. As both were impaired, it is difficult to determine whether the deficit was due to encoding or retrieval. Previous study by Mueller et al. [70] showing that proper noun recall in delayed but not immediate NR is associated with preclinical AD suggests that retrieval is the main factor affected in proper noun recall, however it is important to note that our sample did have biomarker-confirmed AD pathology and may have included other etiologies including vascular dementia.

Mean Yngve depth, a measure of syntactic complexity based on hierarchical relationships between clauses in produced speech, was found to be significantly shallower in picture descriptions of the Cookie Theft image produced by MCI participants. Picture description tasks elicit hierarchical language as participants describe the relationships between objects in a picture or photograph. The Cookie Theft picture in particular contains many relationships between objects, with for example the boy engaging with both the cookie jar and the stool and the man both washing the dishes and not attending to them resulting in them overflowing. The reduction of mean Yngve depth in MCI may be a result of a reduced focus on how objects in a scene relate to each other, instead resulting in simpler descriptions of these objects. Previous studies have shown that cognitive impairment affects mean Yngve depth in NR tasks [19,71], but PD tasks have not been previously shown to elicit decreased syntactic complexity. In our sample, mean Yngve depth did not appear to differ between groups for either NR task but did differ in the PD task. It is possible that the updated Cookie Theft image utilized in our study [34] encouraged either increased interrelated descriptions in healthy controls or a decrease in MCI participants. Additionally, the significant differences were only observed in ANOVAs and depth did not significantly correlate with any individual cognitive assessment (**Figure 2**). While this could be explained due to other mediating effects including age, it is also possible that Yngve depth captures differences not identified in any of the assessments used in our correlations. The PD task also prompted decreased word lengths, Coleman-Liau indices, and noun counts in MCI participants, indicating less precise and simpler language in this task, with this finding supported by previous studies [14,72].

### Use of speech behavior to predict impairment status

After performing feature selection on the entire group of available features, a random forest utilizing the 16 selected features produced a model with an AUC of 0.828 and a DOR of 13.52. While specificity was excellent (94.59%), sensitivity for detecting impairment was only 43.59%. Nonetheless, if the task is to screen for impairment, the use of speech-based markers alone perform well at reducing numbers of non-impaired individuals. When only the top two post-pause features are used, only marginal decreases in sensitivity and specificity are observed. With high specificity and AUC, the results of this model suggest that post-pause metrics of latency and lexical frequency would be an excellent addition to any machine learning model that utilizes speech behavior and seeks to categorize healthy individuals in some capacity, but the low sensitivity precludes their use as a diagnostic tool.

### Limitations

The main limitations of this study were the limited sample size, unequal sample size between impaired and non-impaired groups, the racial and ethnic makeup of the sample being primarily non-Hispanic White, and the lack of biomarker data available in this early sample. As MCI is a heterogeneous disorder, it is possible that separation of MCI subtypes (e.g., MCI due to AD, MCI due to Lewy body or vascular etiologies) will lead to more detailed characterization using neurobehavioral markers such as speech. Additional recruitment and biomarker examination of existing and new participants is ongoing, with increased recruitment efforts targeting impaired as well as racially and ethnically diverse populations.

We also observed age differences between groups. Instead of reducing our healthy control sample to minimize age effects through age-matching, we chose to instead include age as a covariate within our analyses due to the small size of the impaired group. Our future research will incorporate age-matching, as we aim to recruit a sufficient number of biomarker-confirmed impaired participants to support an evenly sized control group. Additionally, some significant measures exhibited standard deviations that exceeded their means, indicating a high degree of variability or skewed distributions for these measures; however, these typically occurred only when all tasks were considered. Thus, this phenomenon is not entirely unexpected as each task was shown to perform differently with respect to each measure and between groups. The only significant task-specific measure with this property was the increased post-pause adverb usage in the free response task.

In this study, only a handful of fillers were examined (“uh”, “um”, and “er”). While many other fillers exist including “you know”, “like”, “okay”, and “right”, particularly among younger individuals, we opted to not include these filler words in our analyses as these were both relatively uncommon in our sample as well as difficult to code in automatic processing; each of these additional filler words can also appear as normal non-filler words, thus requiring manual classification for each of these instances. However, it is possible that the inclusion of these more uncommon filler words would have revealed additional differences between groups. In addition, while the filler “er” was examined in all analyses, no significant differences were found between groups for the usage of this filler nor for words following it. As a result, it is not referenced elsewhere in this paper. Further, this study only examined English-speaking participants: these findings may not generalize to other languages, particularly in the type of frequency of filler words. Nonetheless, this study identified compelling patterns in post-pause speech behavior between MCI and HC in English-speaking populations, and future studies will examine non-native English speakers as well as Spanish speakers to determine whether this behavior translates to other languages and contexts.

## CONCLUSIONS

The increase of post-pause lexical frequency in the absence of task demands and post-filler latency in tasks with high difficulty were observed in MCI participants, with both able to accurately predict impairment status with an AUC of 79.1%. Future research will examine likely causes of pause production, comparing lexical or word-finding pauses with those driven by task demands or cognitive load, as well as neural correlates of speech degradation.

## Supporting information

Table of Features

## ACKNOWLEDGEMENTS

The authors have no acknowledgements to report.

## DATA AVAILABILITY

The data supporting the findings of this study are available on request from the corresponding author.

## REFERENCES

[1] Forbes-McKay KE, Venneri A (2005) Detecting subtle spontaneous language decline in early Alzheimer’s disease with a picture description task. Neurol Sci 26, 243–254.

[2] Ahmed S, Haigh A-MF, de Jager CA, Garrard P (2013) Connected speech as a marker of disease progression in autopsy-proven Alzheimer’s disease. Brain 136, 3727–3737.

[3] Kavé G, Goral M (2016) Word retrieval in picture descriptions produced by individuals with Alzheimer’s disease. J Clin Exp Neuropsychol 38, 958–966.

[4] Fraser KC, Meltzer JA, Rudzicz F (2015) Linguistic features identify Alzheimer’s disease in narrative speech. Journal of Alzheimer’s Disease 49, 407–422.

[5] Yamada Y, Shinkawa K, Nemoto M, Ota M, Nemoto K, Arai T (2022) Speech and language characteristics differentiate Alzheimer’s disease and dementia with Lewy bodies. Alz & Dem Diag Ass & Dis Mo 14,.

[6] Yuan J, Cai X, Bian Y, Ye Z, Church K (2021) Pauses for Detection of Alzheimer’s Disease. Front Comput Sci 0,.

[7] Boschi V, Catricalà E, Consonni M, Chesi C, Moro A, Cappa SF (2017) Connected Speech in Neurodegenerative Language Disorders: A Review. Frontiers in Psychology 8, 269.

[8] Petersen RC (2016) Mild Cognitive Impairment. Continuum (Minneap Minn) 22, 404–418.

[9] Themistocleous C, Eckerström M, Kokkinakis D (2020) Voice quality and speech fluency distinguish individuals with Mild Cognitive Impairment from Healthy Controls. PLOS ONE 15, e0236009.

[10] Mueller KD, Koscik RL, Hermann BP, Johnson SC, Turkstra LS (2018) Declines in connected language are associated with very early mild cognitive impairment: Results from the Wisconsin Registry for Alzheimer’s Prevention. Frontiers in Aging Neuroscience 9, 1–14.

[11] Goodglass H, Kaplan E, Weintraub S (2001) BDAE: The boston diagnostic aphasia examination, Lippincott Williams & Wilkins Philadelphia, PA.

[12] Wechsler D (1987) Wechsler memory scale-revised. The psychological corporation. New York.

[13] Craft S, Newcomer J, Kanne S, Dagogo-Jack S, Cryer P, Sheline Y, Luby J, Dagogo-Jack A, Alderson A (1996) Memory improvement following induced hyperinsulinemia in alzheimer’s disease. Neurobiology of Aging 17, 123–130.

[14] Beltrami D, Gagliardi G, Rossini Favretti R, Ghidoni E, Tamburini F, Calzà L (2018) Speech Analysis by Natural Language Processing Techniques: A Possible Tool for Very Early Detection of Cognitive Decline? Front Aging Neurosci 0,.

[15] Roark B, Mitchell M, Hollingshead K (2007) Syntactic complexity measures for detecting mild cognitive impairment. In Proceedings of the Workshop on BioNLP 2007 Biological, Translational, and Clinical Language Processing - BioNLP ’07 Association for Computational Linguistics, Prague, Czech Republic, p. 1.

[16] Clarke N, Barrick TR, Garrard P (2021) A Comparison of Connected Speech Tasks for Detecting Early Alzheimer’s Disease and Mild Cognitive Impairment Using Natural Language Processing and Machine Learning. Frontiers in Computer Science 3,.

[17] Taler V, Kousaie S, Sheppard C (635775264000000000) Lexical access in mild cognitive impairment. The Mental Lexicon 10, 271–285.

[18] Fraser KC, Lundholm Fors K, Eckerström M, Öhman F, Kokkinakis D (2019) Predicting MCI Status From Multimodal Language Data Using Cascaded Classifiers. Frontiers in Aging Neuroscience 11, 1–18.

[19] Fraser KC, Meltzer JA, Graham NL, Leonard C, Hirst G, Black SE, Rochon E (2014) Automated classification of primary progressive aphasia subtypes from narrative speech transcripts. Cortex 55, 43–60.

[20] Meteyard L, Quain E, Patterson K (2014) Ever decreasing circles: Speech production in semantic dementia. Cortex 55, 17–29.

[21] Goldman-Eisler F (1968) Psycholinguistics: Experiments in spontaneous speech.

[22] Gilden D, Mezaraups T (2022) Laws for pauses. Journal of Experimental Psychology: Learning, Memory, and Cognition 48, 142–158.

[23] Womack K, McCoy W, Ovesdotter Alm C, Calvelli C, Pelz JB, Shi P, Haake A (2012) Disfluencies as Extra-Propositional Indicators of Cognitive Processing. In Proceedings of the Workshop on Extra-Propositional Aspects of Meaning in Computational Linguistics, Morante R, Sporleder C, eds. Association for Computational Linguistics, Jeju, Republic of Korea, pp. 1–9.

[24] Clark HH, Fox Tree JE (2002) Using uh and um in spontaneous speaking. Cognition 84, 73–111.

[25] Shriberg E (2001) To ‘errrr’ is human: ecology and acoustics of speech disfluencies. Journal of the International Phonetic Association 31, 153–169.

[26] Toth L, Hoffmann I, Gosztolya G, Vincze V, Szatloczki G, Banreti Z, Pakaski M, Kalman J (2017) A Speech Recognition-based Solution for the Automatic Detection of Mild Cognitive Impairment from Spontaneous Speech. Current Alzheimer Research 14, 130–138.

[27] Vincze V, Szatlóczki G, Tóth L, Gosztolya G, Pákáski M, Hoffmann I, Kálmán J (2020) Telltale silence: temporal speech parameters discriminate between prodromal dementia and mild Alzheimer’s disease. Clinical Linguistics & Phonetics 00, 1–16.

[28] Egas-López JV, Balogh R, Imre N, Hoffmann I, Szabó MK, Tóth L, Pákáski M, Kálmán J, Gosztolya G (2022) Automatic screening of mild cognitive impairment and Alzheimer’s disease by means of posterior-thresholding hesitation representation. Computer Speech & Language 101377.

[29] Wang T, Hong Y, Wang Q, Su R, Ng M, Xu J, Yan N (2021) Identification of Mild Cognitive Impairment Among Chinese Based on Multiple Spoken Tasks. Journal of Alzheimer’s Disease 82, 1–20.

[30] Besser LM, Chrisphonte S, Kleiman MJ, O’Shea D, Rosenfeld A, Tolea M, Galvin JE (2023) The Healthy Brain Initiative (HBI): A prospective cohort study protocol. PLOS ONE 18, e0293634.

[31] Morris JC (1993) The Clinical Dementia Rating (CDR): Current version and scoring rules. Neurology 43, 2412–2412.

[32] Weintraub S, Besser L, Dodge HH, Teylan M, Ferris S, Goldstein FC, Giordani B, Kramer J, Loewenstein D, Marson D, Mungas D, Salmon D, Welsh-Bohmer K, Zhou XH, Shirk SD, Atri A, Kukull WA, Phelps C, Morris JC (2018) Version 3 of the Alzheimer Disease Centers’ Neuropsychological Test Battery in the Uniform Data Set (UDS). Alzheimer Disease and Associated Disorders 32, 10–17.

[33] Bolognani SAP, Miranda MC, Martins M, Rzezak P, Bueno OFA, de Camargo CHP, Pompeia S (2015) Development of alternative versions of the Logical Memory subtest of the WMS-R for use in Brazil. Dement Neuropsychol 9, 136–148.

[34] Berube S, Nonnemacher J, Demsky C, Glenn S, Saxena S, Wright A, Tippett DC, Hillis AE (2019) Stealing Cookies in the Twenty-First Century: Measures of Spoken Narrative in Healthy Versus Speakers With Aphasia. Am J Speech Lang Pathol 28, 321–329.

[35] Heuer S (2016) The influence of image characteristics on image recognition: a comparison of photographs and line drawings. Aphasiology 30, 943–961.

[36] Hux K, Frodsham K (2023) Speech and Language Characteristics of Neurologically Healthy Adults When Describing the Modern Cookie Theft Picture: Mixing the New With the Old. Am J Speech Lang Pathol 32, 1110–1130.

[37] Radford A, Kim JW, Xu T, Brockman G, McLeavey C, Sutskever I (2022) Robust Speech Recognition via Large-Scale Weak Supervision.

[38] Robert J, Webbie M, Jean-philippe S, Ramdasan A, Lee C, Campbell J, McFarland R, McMellen J, Orby P (2023) PyDub.

[39] Bittner R, Humphrey EJ, Bello J (2016) PySOX: Leveraging the Audio Signal Processing Power of SOX in Python, Proceedings of the 17th International Society for Music Information Retrieval Conference Late Breaking and Demo Papers, New York, NY, USA.

[40] Honnibal M, Montani I, Van Landeghem S, Boyd A (2020) spaCy: Industrial-strength Natural Language Processing in Python.

[41] Speer R (2022) rspeer/wordfreq: v3.0.

[42] Hansen L, Olsen LR, Enevoldsen K (2023) TextDescriptives: A Python package for calculating alarge variety of metrics from text. JOSS 8, 5153.

[43] Coleman M, Liau TL (1975) A computer readability formula designed for machine scoring. Journal of Applied Psychology 60, 283–284.

[44] Brysbaert M, New B (2009) Moving beyond Kucera and Francis: a critical evaluation of current word frequency norms and the introduction of a new and improved word frequency measure for American English. Behav Res Methods 41, 977–990.

[45] Yngve VH (1960) A Model and an Hypothesis for Language Structure. Proceedings of the American Philosophical Society 104, 444–466.

[46] Covington MA, McFall JD (2010) Cutting the Gordian Knot: The Moving-Average Type–Token Ratio (MATTR). Journal of Quantitative Linguistics 17, 94–100.

[47] McCarthy PM, Jarvis S (2010) MTLD, vocd-D, and HD-D: A validation study of sophisticated approaches to lexical diversity assessment. Behavior Research Methods 42, 381–392.

[48] Bortfeld H, Leon SD, Bloom JE, Schober MF, Brennan SE (2001) Disfluency rates in conversation: effects of age, relationship, topic, role, and gender. Lang Speech 44, 123–147.

[49] Vallat R (2018) Pingouin: statistics in Python. JOSS 3, 1026.

[50] Nasreddine ZS, Phillips NA, Bédirian V, Charbonneau S, Whitehead V, Collin I, Cummings JL, Chertkow H (2005) The Montreal Cognitive Assessment, MoCA: A brief screening tool for mild cognitive impairment. Journal of the American Geriatrics Society 53, 695–699.

[51] Cahn-Hidalgo D, Benabou R, Kewin S (2019) Validity, Reliability, and Psychometric properties of Cognivue®, a quantitative assessment of cognitive impairment. The American Journal of Geriatric Psychiatry 27, S212.

[52] Brandt J (1991) The hopkins verbal learning test: Development of a new memory test with six equivalent forms. Clinical Neuropsychologist 5, 125–142.

[53] Galvin JE, Tolea MI, Moore C, Chrisphonte S (2020) The Number Symbol Coding Task: A brief measure of executive function to detect dementia and cognitive impairment. PLOS ONE 15, e0242233.

[54] Kleiman MJ, Galvin JE (2021) The Vulnerability Index: A weighted measure of dementia and cognitive impairment risk. Alzheimer’s & Dementia: Diagnosis, Assessment & Disease Monitoring 13, e12249.

[55] Galvin JE, Kleiman MJ, Chrisphonte S, Cohen I, Disla S, Galvin CB, Greenfield KK, Moore C, Rawn S, Riccio ML, Rosenfeld A, Simon J, Walker M, Tolea MI (2021) The Resilience Index: A Quantifiable Measure of Brain Health and Risk of Cognitive Impairment and Dementia. Journal of Alzheimer’s Disease 84, 1729–1746.

[56] Ke G, Meng Q, Finley T, Wang T, Chen W, Ma W, Ye Q, Liu T-Y (2017) LightGBM: A Highly Efficient Gradient Boosting Decision Tree. In Advances in Neural Information Processing Systems Curran Associates, Inc.

[57] Pedregosa F, Varoquaux G, Gramfort A, Michel V, Thirion B, Grisel O, Blondel M, Prettenhofer P, Weiss R, Dubourg V, Vanderplas J, Passos A, Cournapeau D, Brucher M, Perrot M, Duchesnay E (2011) Scikit-learn: Machine Learning in Python. Journal of Machine Learning Research 12, 2825– 2830.

[58] Keany E (2020) BorutaShap: A wrapper feature selection method which combines the Boruta feature selection algorithm with Shapley values.

[59] Akiba T, Sano S, Yanase T, Ohta T, Koyama M (2019) Optuna: A Next-generation Hyperparameter Optimization Framework. In Proceedings of the 25th ACM SIGKDD International Conference on Knowledge Discovery & Data Mining Association for Computing Machinery, New York, NY, USA, pp. 2623–2631.

[60] Lee J (2019) Task Complexity, Cognitive Load, and L1 Speech. Applied Linguistics 40, 506–539.

[61] Müller C, Großmann-Hutter B, Jameson A, Rummer R, Wittig F* (2001) Recognizing Time Pressure and Cognitive Load on the Basis of Speech: An Experimental Study. In User Modeling 2001, Bauer M, Gmytrasiewicz PJ, Vassileva J, eds. Springer, Berlin, Heidelberg, pp. 24–33.

[62] Swerts M (1998) Filled pauses as markers of discourse structure. Journal of Pragmatics 30, 485–496.

[63] Degand L, Gilquin G, Meurant L, Simon AC (2019) Fluency and Disfluency Across Languages and Language Varieties, Presses universitaires de Louvain.

[64] Earles JL, Kersten AW, Berlin Mas B, Miccio DM (2004) Aging and Memory for Self-Performed Tasks: Effects of Task Difficulty and Time Pressure. The Journals of Gerontology: Series B 59, P285–P293.

[65] Libby LA, Hannula DE, Ranganath C (2014) Medial temporal lobe coding of item and spatial information during relational binding in working memory. J Neurosci 34, 14233–14242.

[66] Kühn S, Gallinat J (2014) Segregating cognitive functions within hippocampal formation: a quantitative meta-analysis on spatial navigation and episodic memory. Hum Brain Mapp 35, 1129–1142.

[67] Russo M, Bendazzoli C, Defrancq B (2017) Making Way in Corpus-based Interpreting Studies, Springer.

[68] Kleiman MJ, Barenholtz E, Galvin JE (2021) Screening for Early-Stage Alzheimer’s Disease Using Optimized Feature Sets and Machine Learning. Journal of Alzheimer’s Disease 81, 355–366.

[69] Yuan J, Bian Y, Cai X, Huang J, Ye Z, Church K (2020) Disfluencies and Fine-Tuning Pre-Trained Language Models for Detection of Alzheimer’s Disease. In Interspeech 2020 ISCA, pp. 2162–2166.

[70] Mueller KD, Koscik RL, Du L, Bruno D, Jonaitis EM, Koscik AZ, Christian BT, Betthauser TJ, Chin NA, Hermann BP, Johnson SC (2020) Proper names from story recall are associated with beta-amyloid in cognitively unimpaired adults at risk for Alzheimer’s disease. Cortex 131, 137–150.

[71] Roark B, Mitchell M, Hosom J-P, Hollingshead K, Kaye J (2011) Spoken Language Derived Measures for Detecting Mild Cognitive Impairment. IEEE Transactions on Audio, Speech, and Language Processing 19, 2081–2090.

[72] Mueller KD, Hermann B, Mecollari J, Turkstra LS (2018) Connected speech and language in mild cognitive impairment and Alzheimer’s disease: A review of picture description tasks. Journal of Clinical and Experimental Neuropsychology 40, 917–939.

