## Supplementary material for "High frequency post-pause word choices and task-dependent speech behavior characterize connected speech in individuals with mild cognitive impairment": Table of Features

| Feature | Description |
| --- | --- |
| Total pause count | Total count of all pauses (both filled and unfilled) |
| Total filled pause count | Total count of all filled pauses |
| Total unfilled pause count | Total count of all unfilled pauses |
| Total “um” filler count | Total count of all “um” fillers |
| Total “uh” filler count | Total count of all “uh” fillers |
| Total “er” filler count | Total count of all “er” fillers (not used) |
| Mean pause duration | Mean duration of all pauses |
| Total word count | Total number of words produced |
| Moving-average type token ratio (MATTR) | Measure of lexical diversity; type token ratio using a moving window of 20 words, unless sample contains less than 20 words |
| Measure of textual lexical diversity (MTLD) | Measure of lexical diversity; calculates type token ratios based on variable windows and both forwards and reversed |
| Mean SUBTLEX lexical frequency | Measure of lexical frequency developed by Brysbaert et al. using a corpus of American English subtitles |
| Mean *wordfreq* lexical frequency | Measure of lexical frequency combining the SUBTLEX corpus with additional web-based sources including Wikipedia, news articles, books, and social media |
| Mean word length | Mean length of all words produced |
| Mean sentence length | Mean length of all sentences produced, determined via period placement from OpenAI’s Whisper |
| Flesch-Kincaid grade level | Readability index; uses word count, sentence count, and syllable count |
| Gunning Fog index | Readability index; uses a measure of complex words (three or more syllables) in addition to total word and sentence counts |
| Coleman-Liau Index | Readability index; uses letter count in addition to word and sentence counts to improve ease of calculation |
| Mean Yngve Depth | Measure of syntactic complexity; quantification of a sentence’s hierarchical structure using the depth of syntactic trees |
| Noun ratio | Ratio of nouns to total words |
| Verb ratio | Ratio of verbs to total words |
| Adjective ratio | Ratio of adjectives to total words |
| Pronoun ratio | Ratio of pronouns to total words |
| Adverb ratio | Ratio of adverbs to total words |
| Proper noun ratio | Ratio of proper nouns to total words |
| Open-class word ratio | Ratio of open-class words to total words |
| Closed-class word ratio | Ratio of closed-class words to total words |
| Closed-to-open-class ratio | Ratio of closed-class to open-class words |
| Noun-to-verb ratio | Ratio of total nouns to total verbs |
| Pronoun-to-noun ratio | Ratio of total pronouns to total noun |
| Adjective-to-verb ratio | Ratio of total adjectives to total verbs |
| Mean post-pause latency | Mean latency between pauses and next word |
| Post-pause features:  *The following features are calculated for each of filled pauses, unfilled pauses, “um” fillers, and “uh” fillers* | |
| 4-word mean post-pause SUBTLEX ranking | Mean SUBTLEX rankings for the 4 words following pauses |
| Single word post-pause SUBTLEX ranking | Mean SUBTLEX rankings for the word following pauses |
| 4-word mean post-pause *wordfreq* ranking | Mean *wordfreq* rankings for the 4 words following pauses |
| Single word post-pause *wordfreq* ranking | Mean *wordfreq* rankings for the word following pauses |
| 4-word mean post-pause noun count | Number of nouns for the 4 words following pauses |
| Single word post-pause noun count | Number of nouns for the word following pauses |
| 4-word mean post-pause verb count | Number of verbs for the 4 words following pauses |
| Single word post-pause verb count | Number of verbs for the word following pauses |
| 4-word mean post-pause adjective count | Number of adjectives for the 4 words following pauses |
| Single word post-pause adjective count | Number of adjectives for the word following pauses |
| 4-word mean post-pause pronoun count | Number of pronouns for the 4 words following pauses |
| Single word post-pause pronoun count | Number of pronouns for the word following pauses |
| 4-word mean post-pause adverb count | Number of adverbs for the 4 words following pauses |
| Single word post-pause adverb count | Number of adverbs for the word following pauses |
| 4-word mean post-pause proper noun count | Number of proper nouns for the 4 words following pauses |
| Single word post-pause proper noun count | Number of proper nouns for the word following pauses |
| 4-word mean post-pause open-class count | Number of open-class words for the 4 words following pauses |
| Single word post-pause open-class count | Number of open-class words for the word following pauses |
| 4-word mean post-pause closed-class count | Number of closed-class words for the 4 words following pauses |
| Single word post-pause closed-class count | Number of closed-class words for the word following pauses |
